# Clinical profile and factors associated with COVID-19 in Cameroon: a prospective cohort study

**DOI:** 10.1101/2021.02.19.21252071

**Authors:** Nicole Fouda Mbarga, Epee Emilienne, Marcel Mbarga, Patrick Ouamba, Herwin Nanda, Aristide Kengni, Guekeme Joseph, Justin Eyong, Sylvie Tossoukpe, Noumedem Sauvia Sosso, Ngono Ngono Engelbert, Mbala Ntsama Lazare, Bonyomo Landry, Tchatchoua Patrick, Noel Vogue, Steve Metomb, Franck Ale, Moussa Ousman, Dorian Job, Moussi Charlotte, Modeste Tamakloe, Jessica E. Haberer, Ndesoh Atanga, Gregory Halle-Ekane, Yap Boum

## Abstract

**Objectives:** This study explores the clinical profiles and factors associated with COVID-19 in Cameroon.

**Research design and methods:** In this prospective cohort study, we followed patients admitted for suspicion of COVID-19 at Djoungolo Hospital between 01^st^ April and 31^st^ July 2020. Patients were categorised by age groups and disease severity: mild (symptomatic without clinical signs of pneumonia pneumonia), moderate (with clinical signs of pneumonia without respiratory distress) and severe cases (clinical signs of pneumonia and respiratory distress not requiring invasive ventilation). Demographic information and clinical features were summarised. Multivariable analysis was performed to predict risk.

**Results:** A total of 323 patients were admitted during the study period; 262 were confirmed cases of COVID-19 by Polymerase Chain Reaction (PCR). Among the confirmed cases, the male group aged 40 to 49 years (13.9%) was predominant. Disease severity ranged from mild (77%; N=204) to moderate (15%; N=40) to severe (7%; N=18); the case fatality rate was 1% (N=4). Dysgusia (46%; N=111) and hyposmia/anosmia (39%; N=89) were common features of COVID-19. Nearly one-third of patients had comorbidities (29%; N=53), of which hypertension was the most common (20%; N=48). Participation in a mass gathering (OR=5.47; P=0.03) was a risk factor for COVID-19. Age groups 60 to 69 (OR=7.41; P=0.0001), 50 to 59 (OR=4.09; P=0.03), 40 to 49 (OR=4.54; P=0.01), male gender (OR=2.53; P=0.04), diabetes (OR= 4.05; P= 0.01), HIV infection (OR=5.57; P=0.03), lung disease (OR= 6.29; P=0.01), dyspnoea (OR=3.70; P=0.008) and fatigue (OR=3.35; P=0.02) significantly predicted COVID-19 severity.

**Conclusion:** Unlike many high-income settings, most COVID-19 cases in this study were benign with low fatality. Such findings may guide public health decision-making.

## Introduction

In December 2019, COVID-19 was first identified in Wuhan, Capital City of Hubei Province, in China.^1^ COVID-19 is caused by severe acute respiratory syndrome coronavirus 2 (SARS-CoV-2), a novel RNA betacoronavirus.^2^ COVID-19 was declared a public emergency of international concern on 30th January 2020 by the World Health Organisation (WHO) and it became a pandemic on the 11th March 2020, acknowledging the rapid spread of the disease across continents.^3^

In Cameroon, the first case was reported on the 06^th^ March 2020, a traveler who arrived in Cameroon on the 24^th^ February 2020 from France. As of the 17^th^ August 2020, a total of 18118 cases (1177 active cases, 16540 recoveries and 401 deaths [2.2%]) were declared.^4^ This low case fatality rate (CFR) 05 months after declaration of the onset of the epidemic in Cameroon contrasts with an average CFR of 4% globally with in country CFR after the 100^th^ case ranging from 0% in countries like Bahrain to more than 20% in France^5^. This calls for an analysis of the disease pattern, distribution, and clinical characteristics in the African context. However, so far, most evidence on COVID-19 have been described in high-income settings.^6–9^

In Africa, the epidemiology of COVID-19 may differ from that reported in Europe, Asia, and America for a number of reasons. First, like most other African countries, Cameroon suffers from the double burden of infectious diseases (e.g. tuberculosis, HIV/AIDS and malaria) and non-communicable diseases (e.g., sickle cell diseases, cancer, cardiovascular diseases, diabetes, and renal diseases amongst others) ^10^. In addition, confinement was not the hallmark of mitigation efforts in Cameroon.^11^ For these reasons, we expected all these factors to contribute to an escalation of the transmission of COVID-19 in Cameroon. This study was aimed at providing a characterisation of epidemiological profile of cases and risk factors associated with COVID-19 infection at the Djoungolo Hospital thereby providing more knowledge of SARS-Cov-2 transmission in the Cameroonian context.

## Summary Box

### Evidence before this study

Evidence suggests that COVID-19 infection has a wide range of manifestations including olfactory and gustatory symptoms. Findings also imply that the COVID-19 infection is benign in most cases (up to 70%) and fatality varies from country to country, with Italy, Spain and France which experienced very high fatalities when compared with other countries like South Korea and China. Male gender, symptoms like dyspnoea and fatigue, and comorbidities such as diabetes and lung disease are risk factors for COVID-19 severity.

### Added value of this study

The study suggest that HIV infection is also a risk factor for severe COVID-19 infection in Cameroon. The findings imply that COVID-19 is benign for most cases in Cameroon with low lethality. These findings call for enforcement of public health measures on prevention of COVID-19 in elderly patients with co-morbidities.

## Methods

### Ethical oversight

Ethical clearance (Number 2020/09/1294/CE/CNERSH/SP) was sought from the National Ethical Committee of Research for Human Health in Cameroon. Administrative authorisations were also obtained from all authorities concerned.

### Study setting and design

Cameroon, located at the heart of Central Africa, is a country with a population essentially made up of blacks and a younger population when compared to that of many Western countries^12^ with only 2.7% of people aged more than 65 years.^13^ The environment is also distinct with high temperatures and meteorological variabilities.^14^ Moreover, the sociocultural context is unique with strong beliefs in traditional medicine.^15^ Cameroon adopted public health measures which evolved with the trends of the pandemic. On the 17th March 2020, social regulations were enforced in Cameroon: closure of borders, confinement at home from 6PM, and regulations on transportation means (passengers 1m apart).^11^ Towards 13^th^ April 2020, it became compulsory to wear a facial mask.^16^ Specialized COVID-19 treatment centres and laboratories were identified for diagnosis and management of COVID-19 in Cameroon. The Djoungolo Hospital, located in Yaounde, the Centre Region, was one of the six COVID-19 treatment centres dedicated officially for the management of suspected and confirmed cases of COVID-19 by the Ministry of Public Health and was supported by Medecins Sans Frontieres (MSF). We used a prospective cohort study design to follow patients admitted at the Djoungolo Treatment Centre for COVID-19 infection from the 01^st^ April 2020 to the 31^st^ July 2020. At the beginning of the pandemic, COVID-19 patients were discharged from the hospital based on availability of a negative PCR results after 14 days of hospital admission.

### Study participants

We included patients admitted at the Djoungolo Treatment Centre for suspicion of COVID-19 and with a confirmed diagnosis of COVID-19 by PCR according to national protocols proposed by the Ministry of Public Health. The standard WHO case definition of COVID-19 was used as a checklist for admission^17^. Suspected cases who ultimately had no laboratory confirmation and non-cases were excluded. The study population was organised into age groups as follows: 0 to 17, 18 to 29, 30 to 39, 40 to 49, 50 to 59, 60 to 69 and more than 70 years. Patients were categorised by disease severity as mild (i.e. symptomatic without clinical signs of pneumonia pneumonia), moderate (with clinical signs of pneumonia without respiratory distress) and severe cases (clinical signs of pneumonia and respiratory distress not requiring invasive ventilation)^17^. Patients who required invasive ventilation were considered critical and had to be referred to specialised centres when possible. We considered mild and moderate severity levels as benign forms of COVID-19.Patients with more than 80% missing variables were automatically excluded from the analysis.

### Data collection

Socio-demographic and clinical events were recorded on a case-report form (CRF) designed exclusively for all suspected or confirmed cases of COVID-19 patients admitted in Djoungolo Hospital. This CRF was conceived using the forms proposed by the MOH, the WHO and MSF/Epicentre. Sociodemographic variables collected included gender, age, religion, profession, residence, risk of exposure (i.e., recent travel, exposure to health facility, exposure to a traditional healer, and any other form of attendance to a gathering), and contact tracing (i.e., contacts traced and tested with corresponding dates). Clinical events were recorded including symptoms and date of onset, comorbidities, patient itinerary (from onset of symptoms to discharge), complications, diagnostic procedure, test results, and treatment provided.

### Data management and analysis

Continuous variables were expressed as medians and simple ranges, as appropriate. Categorical variables were summarised as counts and percentages. Chi-square tests and Fisher’s exact tests were used for categorical variables. Epidemic curves were constructed with admission dates at the treatment centre. An onset-to-admission curve was constructed by fitting a log-normal distribution to data on both onset and diagnosis dates. Multivariate logistic regression analysis was performed to determine the predictors of COVID-19 disease and disease severity. To predict risk of disease severity, the glmulti package was used in R for automated model selection and multimodel inference^18^. The fit and plausibility of various models^18^ were examined in RStudio, focusing on models which contained none, one, and up to seven (i.e., all) of moderator variables for risk of severity. We used two groups of moderator variables. Group 1 included age groups, gender, comorbidities (i.e., HIV infection, diabetes, lung disease and hypertension). Group 2 included age group, gender and symptoms (i.e., dyspnea, cough, chest pain). We considered models with main effects only. The Akaike information criterion (AIC) ^19^ was used to choose the most performant model. The Akaike weight for a particular model can be regarded as the probability that the model is the best model (in a Kullback-Leibler sense of minimizing the loss of information when approximating full reality by a fitted model) out of all of the models considered.^19^ In other words, the best model was considered to have the lowest AIC with Akaike weights and Kullback-Leibler divergence indicating the highest probability of model accuracy. Data analysis was performed with STATA version 13 and RStudio Version 1.3.1073 for modelling.

### Operational Definitions

Time to hospitalisation: the time interval between the onset of the first symptom and the date of admission.

Length of hospital stay: time interval between the admission date and the date the patient is discharged or dies.

Mass gathering: travel and exposures such as a visit to a church or a mosque, a hospital, a market or

Laboratory confirmation: confirmation for SARS-CoV-2 positive PCR assays is performed in Yaounde by several laboratories in accordance with the protocol established by the WHO^17^.

### Patient and public involvement

Not considered.

## Results

### 1. Patients characteristics

During the study period, a total number of 337 patients were admitted at the Djoungolo Treatment Centre with suspicion for COVID-19, among which 262 (77%) were confirmed cases. Admissions evolved in a sawtooth– like wave, with some peaks in May and June and a major drop in admissions around late June 2020. The number of cases admitted daily ranged from 1 to 9 patients (Figure 1). A total of 26 patients (8%) were excluded from the analysis because of missing data. Among all confirmed cases, there was a male predominance and the most prevalent age category was 40 to 49 years (13.9%) (Figure 2 &3). In the group of severe patients, there was also a male predominance but the most common age groups were 40 to 49 (23%) and 50 to 59 years (23%).

**Figure 1:**
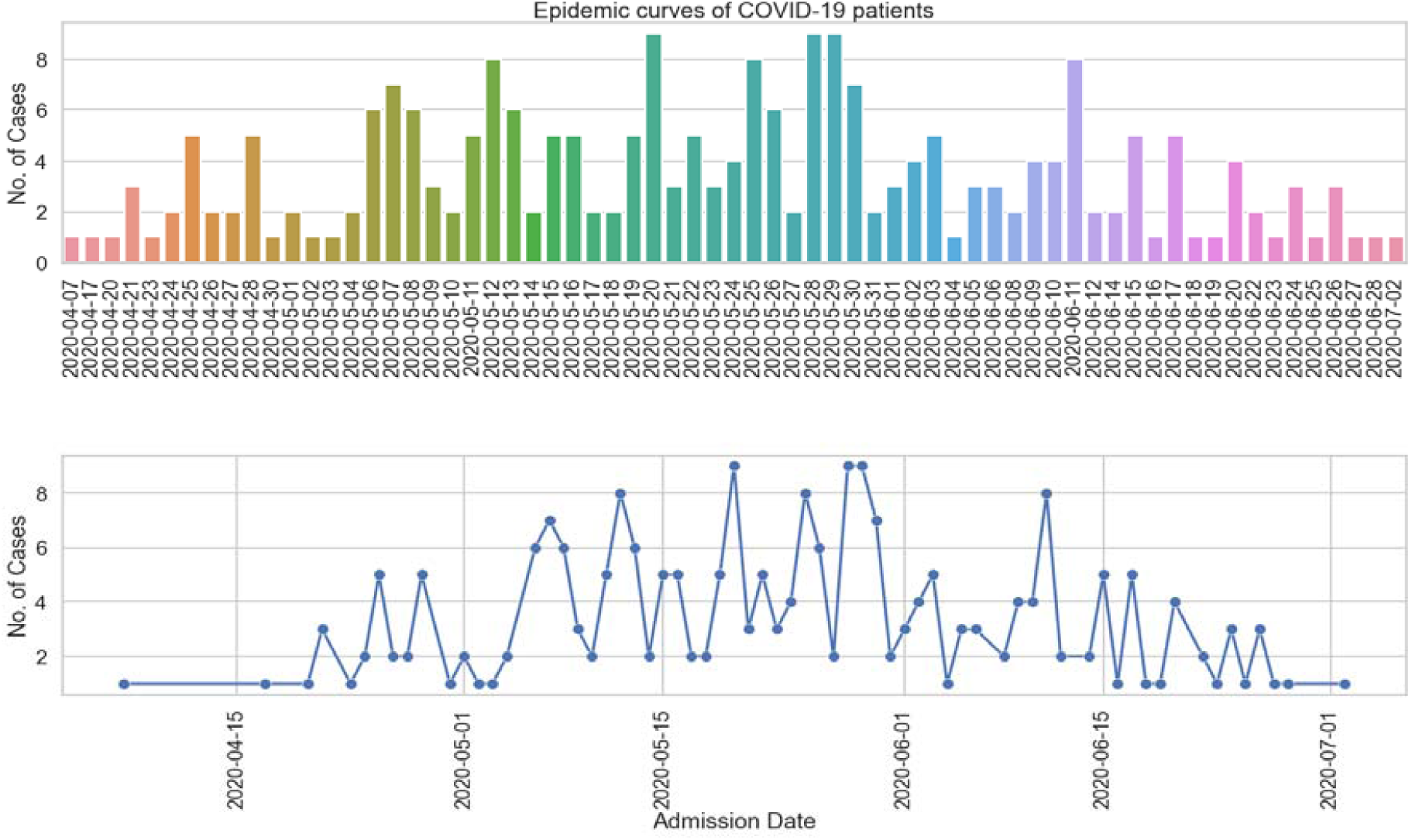
Epidemic curve of Covid19 cases at the Djoungolo treatment centre.

**Figure 2:**
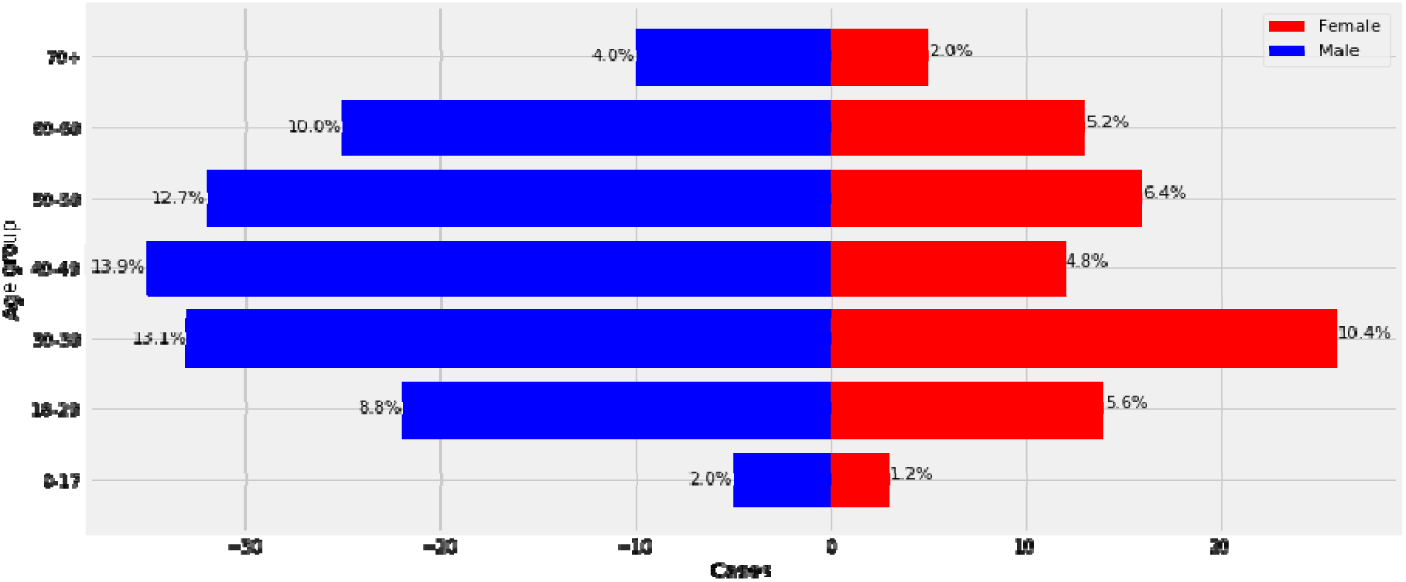
Age/sex pyramid (all cases)

**Figure 3:**
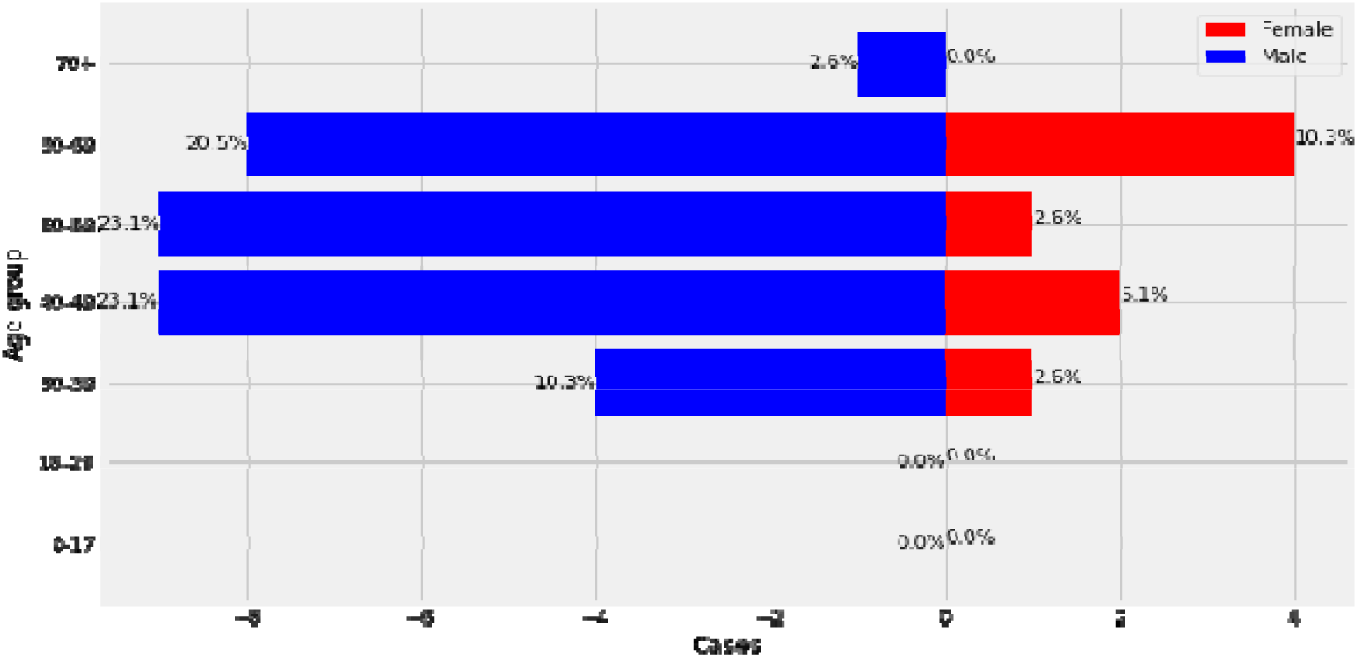
Age/sex pyramid (Severe cases)

### 1. Clinical profile

#### Symptoms and signs

Most patients presented with a cough (61%; N=145), fever (53%; N=125), dyspnoea (52%; N=122), fatigue (52%; N=122), and headaches (50%; N=118). Other common symptoms were dysgusia (46%; N=111), arthralgia or myalgia (43%; N=100), and hyposmia/anosmia (39%; N=89). Nasal discharge (4%; N=9) and sore throat (7%; N=17) were the least common symptoms (Figure 4).

**Figure 4:**
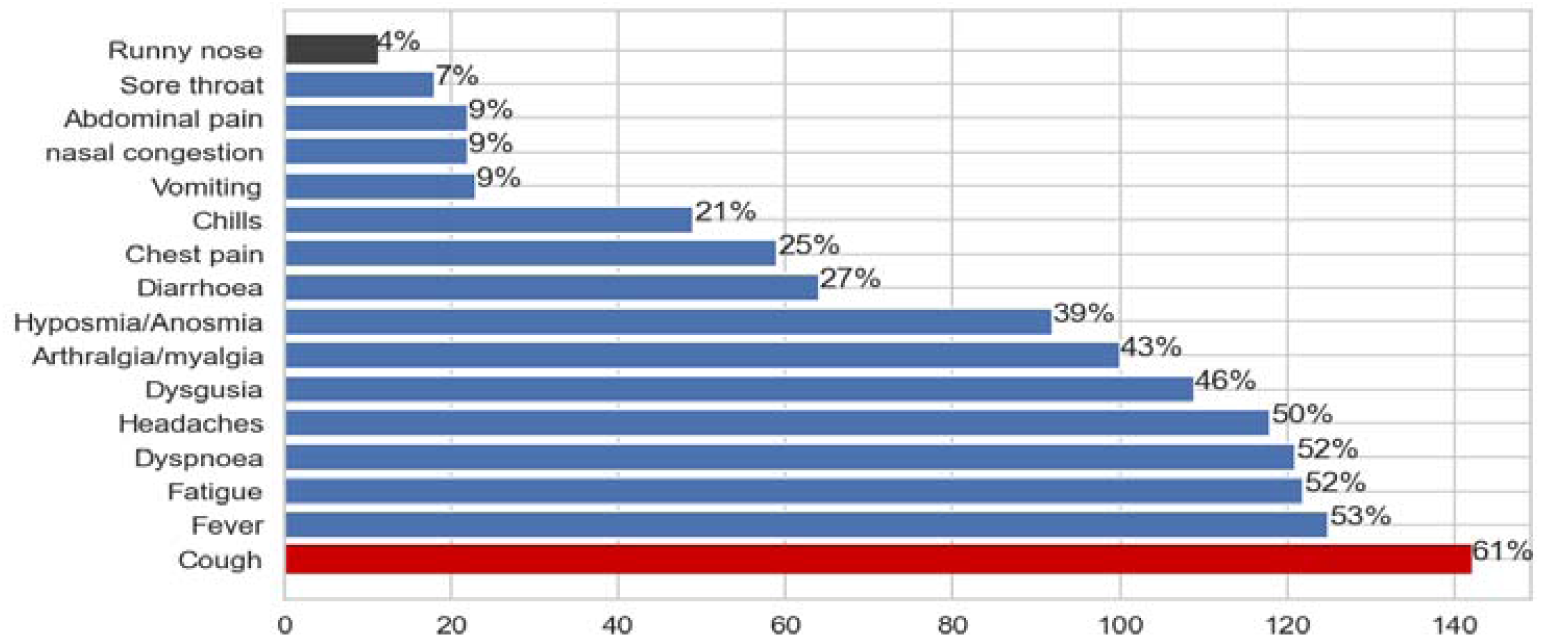
Proportion of symptoms and signs at admission.

#### Severity and comorbidities

Most patients had benign disease (77%; N=204), some patients had severe disease (15%; N=40) and required a supplemental oxygen supply, and a few patients were critical and required a transfer to specialised centres (7%; N=18). However, it is worth mentioning that only 30% of critical patients who required a transfer were finally transferred as a result of overcrowding in sophisticated COVID-19 treatment centres with intensive care units. Four confirmed cases died, yielding a CFR of 1%. Twenty-nine percent (N=187) had at least one comorbidity. The most common comorbidities were hypertension (20%; N=48), other cardiovascular diseases (8%; N=18), diabetes (6%: N=13), and HIV infection (3%; N=7) (Figure 3). Three out of the 04 patients (75%) who died had hypertension; one patient had both hypertension and diabetes. The deceased cases were aged 53, 66, 68, and 73 years

#### Hospital course

The average length of hospitalisation was 15 days (10-26 days). Length of stay was lowest for male children aged 11 months to 17 years and highest for male patients aged more than 70 years (Figure 5). Time to hospitalisation ranged from 0 to 40 days for some patients, but the majority of patients were admitted 07 to 13 days after symptom onset (Figure 6).

**Figure 5:**
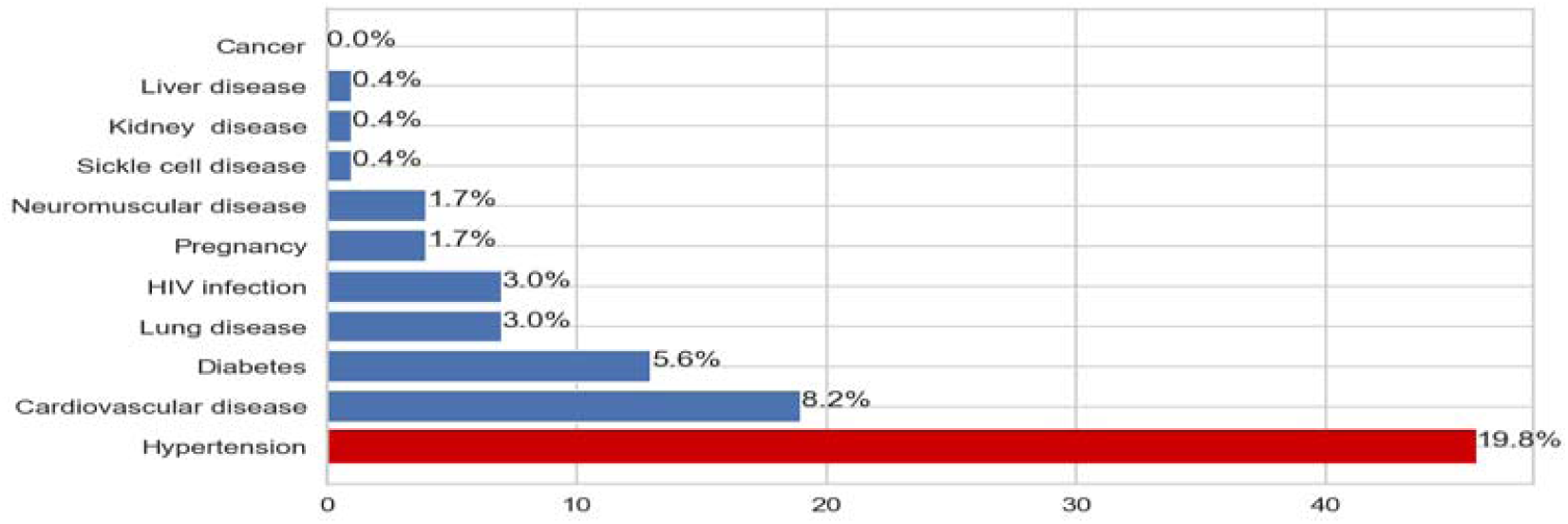
Proportion of comorbidities among admitted patients.

**Figure 6:**
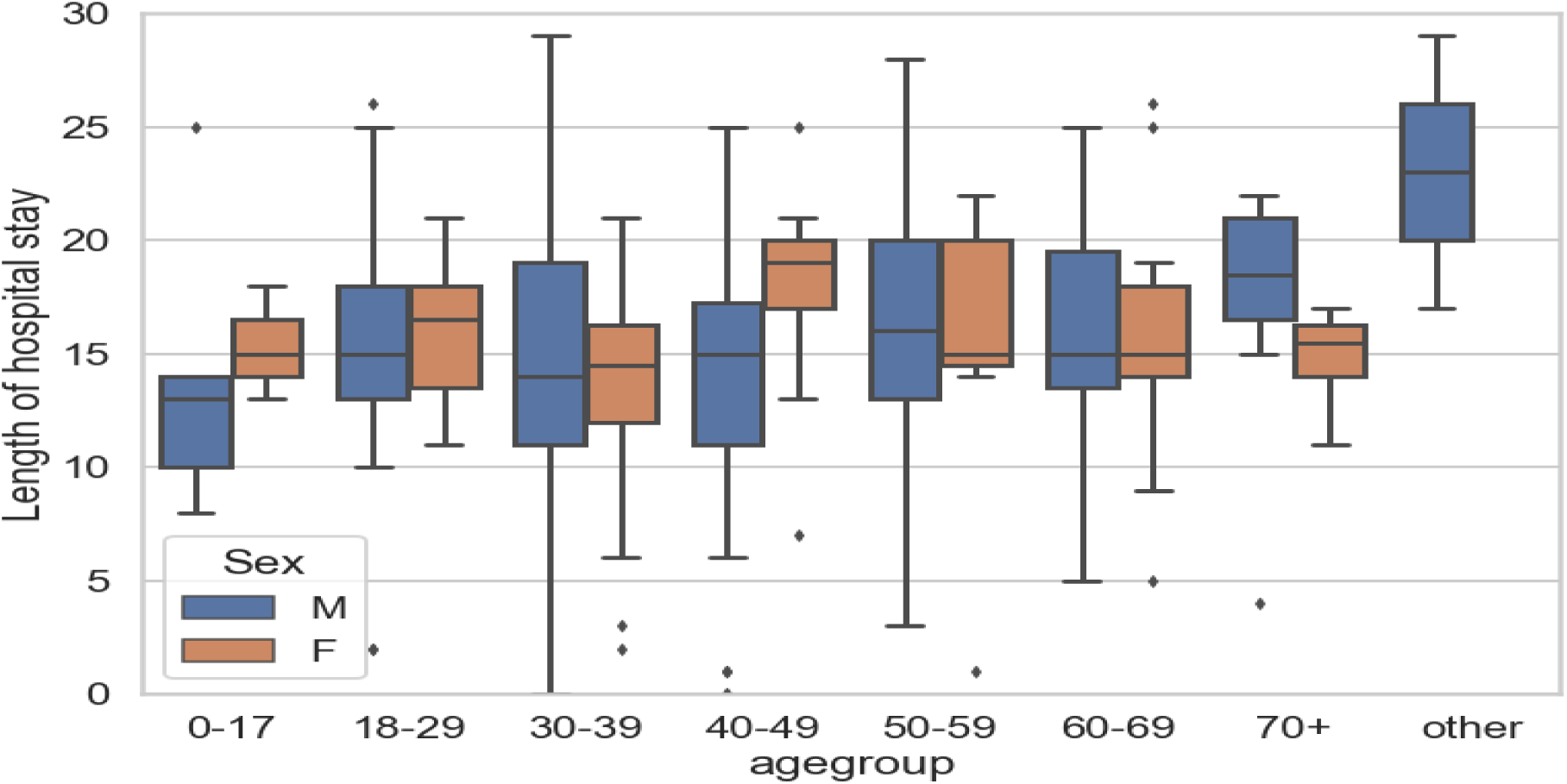
Length of hospital stay by age group.

**Figure 7:**
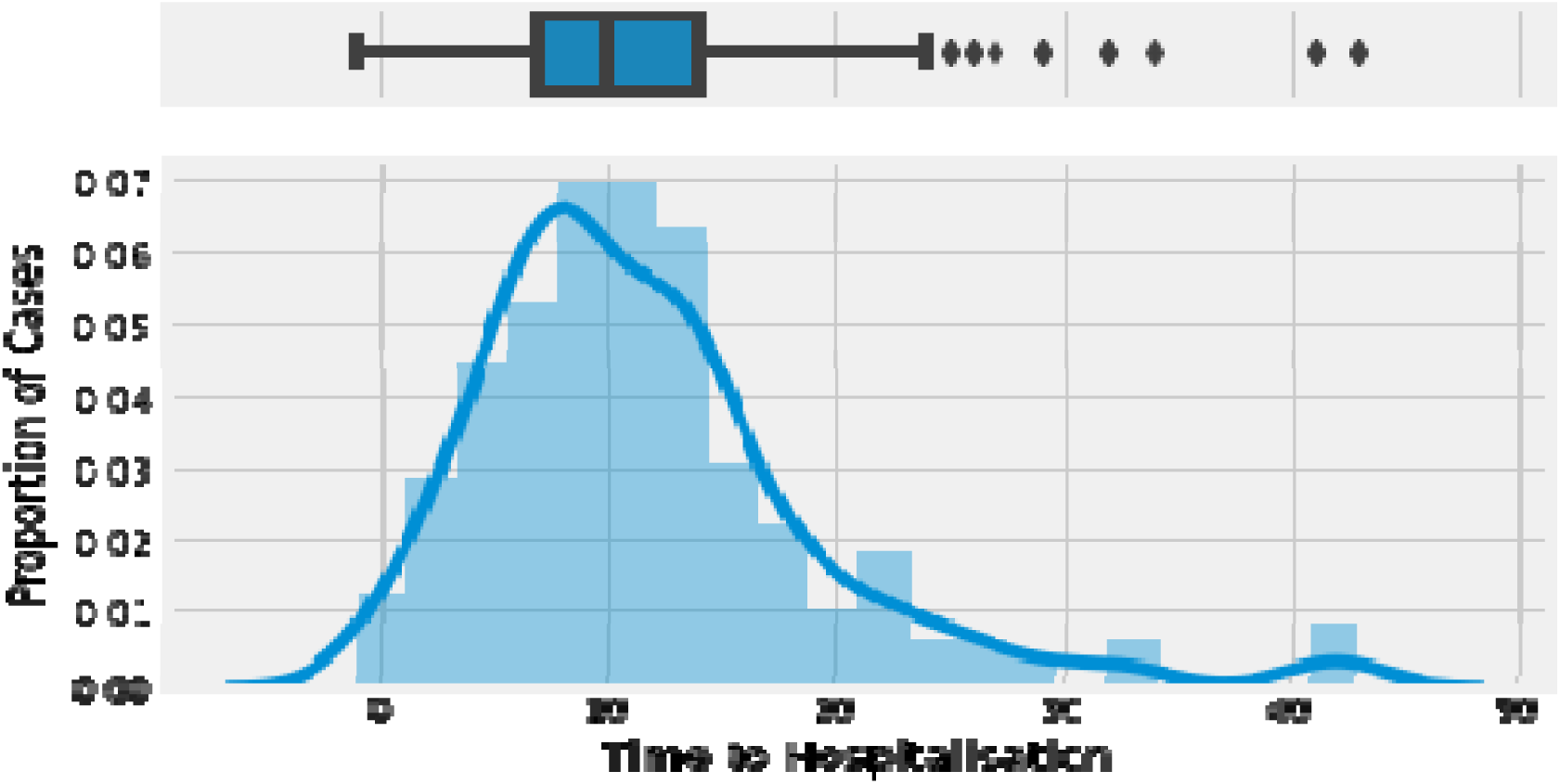
Admission delay curve.

#### Treatment

All patients received a treatment protocol with oral chloroquine, paracetamol, vitamin C, zinc, amoxicillin combined with clavulanic acid, and azithromycin. Depending on severity and comorbidities, some patients received anticoagulants, corticosteroids or intravenous antibiotics. Fifteen percent of confirmed cases underwent non-invasive ventilation (N= 40).

### 1. Risk factors

#### Factors associated COVID-19 diagnosis

We compared groups of confirmed (82%; N=259) cases and non-cases (18%; N= 54). Participation at a mass gathering significantly increased the odds of being diagnosed with COVID-19 (OR=5.47; P=0.03: CI=1.15-25.88). Being aged 30 to 39 (OR= 0.03; P=0.03; CI= 0.01-0.76), a prisoner (OR=0.03; P=0.01; CI=0.01-0.34), retired (OR= 0.03; P=0.01; CI=0.01-0.42), or a student (OR=0.06; P=0.01; CI= 0.01-0.79) were significantly protective factors against COVID-19 in our settings. There was no significant relationship between age (>50 years), male gender, or travel history and a confirmed COVID-19 diagnosis (Table 1).

**Table 1:** Output for multivariable analysis of factors associated with a diagnosis of COVID-19.

|  | OR | Std<br>Error | Z | P value | 95% Confidence interval |  |
| --- | --- | --- | --- | --- | --- | --- |
| 1. Age group |  |  |  |  |  |  |
| 0-17 | Reference |  |  |  |  |  |
| 18-29 | 0.50 | 0.68 | -0.51 | 0.61 | 0.03 | 7.03 |
| 30-39 | **0.03** | **0.05** | **-2.13** | **0.03** | **0.01** | **0.76** |
| 40-49 | 0.08 | 0.13 | -1.56 | 0.11 | 0.01 | 1.91 |
| 50-59 | 0.14 | 0.24 | -1.17 | 0.24 | 0.01 | 3.71 |
| 60-69 | 1.50 | 2.90 | 0.21 | 0.83 | 0.03 | 65.78 |
| 70+ | 0.53 | 1.01 | -0.33 | 0.74 | 0.01 | 22.14 |
| 2. Gender |  |  |  |  |  |  |
| Female | Reference |  |  |  |  |  |
| Male | 0.67 | 0.31 | -0.84 | 0.39 | 0.27 | 1.67 |
| 3. Profession |  |  |  |  |  |  |
| Employed formal<br>(not health profession) | Reference |  |  |  |  |  |
| Employed informal | 0.33 | 0.21 | -1.67 | 0.09 | 0.09 | 1.20 |
| **Health professional** | **0.17** | **0.14** | **-2.03** | **0.04** | **0.03** | **0.94** |
| **Prisoners** | **0.03** | **0.03** | **-2.86** | **0.01** | **0.01** | **0.34** |
| **Retired** | **0.03** | **0.04** | **-2.61** | **0.01** | **0.01** | **0.42** |
| **Student** | **0.06** | **0.08** | **-2.14** | **0.03** | **0.01** | **0.79** |
| 4. Exposure |  |  |  |  |  |  |
| No exposure | **Reference** |  |  |  |  |  |
| **Mass gathering** | **5.47** | **4.33** | **2.15** | **0.03** | **1.15** | **25.88** |
| Travel history | 0.98 | 0.51 | -0.03 | 0.97 | 0.34 | 2.76 |
| 5. History of Smoking |  |  |  |  |  |  |
| No smoking | **Reference** |  |  |  |  |  |
| Smoking | 0.40 | 0.49 | -0.74 | 0.45 | 0.03 | 4.34 |

#### Factors associated with COVID-19 severity

The most important variables for prediction of risk of COVID19 severity were: diabetes, lung disease, HIV infection, male gender, and age groups 40 to 49, 50 to 59, and 60-to 69. In our settings, age groups 60 to 69, 50 to 59, 40 to, male gender, diabetes, HIV infection and lung disease were significant predictors of COVID-19 severity (Table 2). Dyspnoea and fatigue are warning signs of COVID-19 severity (Table 3).

**Table 2:** multivariable analysis of comorbidities associated with COVID-19 severity.

|  | OR | Std Error | t Value | P value | 95% Confidence Interval |  |
| --- | --- | --- | --- | --- | --- | --- |
| Male gender | 2.53 | 0.04 | 1.98 | 0.04* | 1.06 | 6.76 |
| Age group 40 to 49 | 4.54 | 0.05 | 2.37 | 0.01* | 1.52 | 14.63 |
| Age group 50 to 59 | 4.09 | 0.05 | 2.16 | 0.03 * | 1.36 | 13.18 |
| Age group 60 to 69 | 7.41 | 0.06 | 3.56 | 0.0001*** | 2.52 | 23.85 |
| HIV infection | 5.57 | 0.13 | 0.13 | 0.03* | 0.92 | 31.76 |
| Diabetes | 4.05 | 0.09 | 2.53 | 0.01* | 1.12 | 14.15 |
| Lung disease | 6.29 | 0.13 | 0.14 | 0.02* | 0.91 | 44.93 |

\*Level of statistical significance

**Table 3:** multivariable analysis of symptoms associated with COVID-19 severity.

|  | OR | Std error | t value | P- value | 95% Confidence interval |  |
| --- | --- | --- | --- | --- | --- | --- |
| Male gender | 2.75 | 0.47 | 2.12 | 0.008* | 1.12 | 7.4 |
| Age group 40 to 49 | 4.79 | 0.60 | 2.58 | 0.009** | 1.49 | 16.63 |
| Age group 50 to 59 | 3.19 | 0.59 | 1.95 | 0.05 | 1.01 | 10.7 |
| Age group 60 to 69 | 6.65 | 0.59 | 3.13 | 0.001** | 2.07 | 22.3 |
| Fatigue | 3.35 | 0.52 | 2.31 | 0.02* | 1.26 | 10.1 |
| Dyspnoea | 3.70 | 0.49 | 2.64 | 0.008** | 1.46 | 10.45 |
| Abdominal pains | 0.31 | 0.8 | 61.42 | 0.15 | 0.04 | 1.27 |
| Muscle pains | 2.25 | 0.59 | 1.93 | 0.05 | 1.00 | 5.23 |

\*Level of statistical significance

## Discussion

The study sought to describe clinical features of COVID-19 infection and risk factors for patients admitted in the Djoungolo Hospital COVID-19 treatment centre in Cameroon. This study adds to the description of COVID-19 in African hospital settings where there is a huge gap in the literature. Most patients experienced a benign form of COVID-19 infection with low fatality. Being aged 30 to 39 years, being a health professional, a prisoner, retired or being a student appeared as significant protective factors against COVID-19.Mass gathering was a risk factor of COVID-19 transmission and infection. Male gender, older age, comorbidities (i.e., diabetes, HIV, and lung disease) and the presence of fatigue and dyspnoea were predictors of COVID-19 severity.

Analysis of COVID-19 severity and mortality varies across the world. Grant et al reported findings synthesised from studies conducted across 10 Western countries: United Kingdom UK, the United States of America, Singapore, Italy, Australia, Japan, Korea, and the Netherlands.^20^ These authors indicated that 19% of patients hospitalised in these countries required non-invasive ventilation, which is quite close to the value found in our setting.^20^ The CFR was relatively low at the Djoungolo Treatment Centre (1%) and this can be partially explained by standard operating procedures for management at Djoungolo Hospital which required transfer of critical cases to more sophisticated treatment centres. However, due to overcrowding in sophisticated centres, some critical patients were managed at the Djoungolo Hospital and this accounts for the fatalities reported. Nonetheless, the CFR obtained in this study was similar to the Cameroonian National CFR estimated at 2% as of the 07^th^ October 2020^4^. Our CFR was also similar to the average CFR reported in South Korea and Germany, while countries like France and Belgium had relatively high CFR (20 and 16% respectively).^5^ Grant et al found a CFR of 7% in their meta-analysis.^20^ This variability could be due to the difference of age of confirmed cases. As seen in our study, more than 55% of patients in our study were aged less than 50 and the mean age was 44 years as compared to 49 years as reported by Grant et al.^20^

Clinical features of COVID-19 infection in our setting were slightly different from those reported by studies in high-income countries. The Grant et al meta-analysis reported: fever (78% [95% CI 75%-81%] in 138 studies, 21,701 patients; I2 94%), cough (57% [95% CI 54%-60%]; 138 studies, 21,682 patients; I2 94%) and fatigue (31% [95% CI 27%-35%]; 78 studies, 13,385 patients; I2 95%) as the most common symptoms. Fatigue (31%, CI= 27-35%, 31 studies; I2= 95%), dyspnoea (23%, CI=19-28%; 94 studies; I2=97%) and headaches (13%, CI=10-16, 65 studies) had smaller proportions than those reported in our study.^20^ In the same line with Grant et al, gastrointestinal manifestations, sore throat, and nasal discharge were also less common in the current study. In our study, ophthalmic manifestations were not prevalent. Differences in population characteristics (i.e., age, race, genetic make-up or immunity) and even settings (climate and environment) might explain the differences noticed in the findings.

Dysgeusia (46%; N=111) and hyposmia/anosmia (39%; N=89) had higher prevalence in our study when compared with findings reported by Grant et al (i.e., hypogusia 25%, hyposmia 4%).^20^ In a systematic review recently published by Mehraeen et al, 95% of studies had anosmia as a feature of COVID-19 infection^21^. It is worth mentioning that Mehraeen et al and Grant et al had similar methods as they both included evidence from at least 10 Western countries and conducted their searches from December 2019/January 2020 to April 2020. However, Grant et al had no language restrictions and reviewed a wider range of papers. This difference in scope might explain the difference in findings. In general, there is growing evidence that COVID-19 also presents with olfactory as well as gustatory dysfunction, but the pathogenesis is not yet known.^21^ This finding could imply that the current case definition of COVID-19 published by the WHO may need to be updated^22^.

In our study, 29% of patients suffered from a comorbidity, while Morgan et al had twice more patients who had at least 01 comorbidity (40%) in a meta-analysis of COVID-19 and comorbidities. Morgan et al synthesized evidence from Western countries including China, South Korea, Australia, and the USA, which have relatively older populations^23^. This difference in ages might explain the higher prevalence of cases with comorbidities in their settings. They also found the commonest comorbidity to be hypertension and the majority of fatal cases had at least 01 comorbidity (74%), in agreement with our findings. They equally observed that diabetes and respiratory illnesses were prevalent in the deceased^23^, which is not the case in the current study where hypertension was identified as common in the deceased.

The median length of hospital stay was 15 (10 to 27 days) in this study. In a systematic review of length of hospital stay for COVID-19 patients, Rees et al observed that length of hospital stay was generally higher for patients followed up in China. The median range of length of hospital stay ranged from 4 to 53 days in China and 4 to 21 days elsewhere (i.e., Europe, USA and UK).^24^ Most studies included by Rees et al were conducted in China (88%) and specifically in Wuhan. Lengthy hospital stays reported by Rees et al can be explained by inclusion of studies analysing critically ill and old patients (average 68-69 years). In our context, long hospital stays were equally observed in patients aged more than 70 years. The speed at which the last negative test for SARS-CoV2 was obtained also explains lengthy stays in our study. With the reduced testing capacity at the beginning of the epidemic in Cameroon, it was a real challenge to obtain PCR results after 14 days. Additionally, length of hospitalization can vary by the culture of clinical practice, as well as by disease severity and logistical constraints, thus complicating comparisons among international settings.

Unexpectedly, being aged 30 to 39, being a prisoner, retired, or a student were protective against COVID-19 infection in this study. These findings may be explained by the public health measures implemented a few days after the declaration of the COVID-19 epidemic in Cameroon required closures of schools^11^, which might have been protective for students. Moreover, public health measures to fight COVID-19 epidemic in Cameroon also concerned prisoners and included reduced movement among prisons, reinforcement of hygiene measures, and special authorisation of release for some prisoners^11^. These policies might have impacted the circulation of the SARS-CoV-2 virus in prisons. In addition, retired individuals tend to move less than workers^25^ and this might have reduced their exposure to the COVID-19 infection.

Elsewhere, increasing age, male gender, comorbidities (e.g., diabetes, lung disease) have also been associated with COVID-19 severity^26-28,^. In addition, HIV infection appeared as a significant predictor of severity in this study though not reported in Western settings. In Cameroon, HIV prevalence was estimated at 2.9% in 2019, with 75% of HIV infected people knowing their status and only 62% on treatment^29^. Perhaps this burden might explain our findings. The most significant variable for prediction of COVID-19 severity in the current study was age group 60 to 69 and unexpectedly, people aged more than 70 years were not at increased risk of COVID-19 severity in our study. The small representation of patients aged more than 70 years in our sample might explain this finding. In a meta-analysis conducted by Rahman et al, male gender, hypertension, diabetes, fatigue or myalgia, and smoking history were found to be risk factors for COVID-19 severity.^26^ There are some differences in comparison with our results, which do not include smoking, hypertension as risk factors for severity. Studies included in the meta-analysis were conducted in China, perhaps these differences in population characteristics such as smoking history might account for differences in risk factors for severity. For instance, Rahman et al had a proportion of 11% of cigarette smokers compared with 1% in our study population^26^. On the other hand, diabetes and male gender have been consistently reported to potentially increase the risk of suffering from severe form of COVID-19 in other settings.^27–28^

This study also provided evidence that mass gatherings contribute to the spread of COVID-19. Apart from social gatherings, mass gatherings and high density activities in general are associated with generation of income and revenues through formal or informal employment and other businesses.^30^ Cameroon faced a dire dilemma between restricting all mass gatherings to curb COVID-19 while suffering from an economic crisis and choosing to let businesses be open and taking the risk of increasing the spread of COVID-19, which is particularly challenging given its developing status. Due to the effects on the economy, on 30^th^ April 2020, the government relaxed social distancing measures^11^. Restrictions were lifted on opening hours for businesses and even transportation. This change might explain, at least in part the rise in admissions at the Djoungolo Treatment Centre from May 2020 to June 2020.

This study adds to the knowledge gap on COVID-19 in African settings. Despite its original content, our study has limitations inherent to the observational design. Only 50 suspected cases had no evidence of infection with SARS-CoV2, which might have impacted the multivariable analysis of risk of COVID-19 infection. The study was conducted at a single centre, thus generalisability of findings nationwide might not be accurate. Additionally, conducting rigorous research in a context of a novel pandemic was challenging and missing data was common. For instance, incubation periods were not calculated as only 07% had required data. Moreover, in our setting, laboratory and radiological diagnostics could not be systematically performed for patients. Some critical cases were referred to more sophisticated centres as required and this might in part explain the low CFR obtained at the Djoungolo Hospital.

In this study, we described clinical features of COVID-19 infection in a typical COVID-19 treatment centre in Cameroon. The Djoungolo Treatment Centre experienced its highest number of admissions from early May to mid-June. Most patients had a benign form of COVID-19 and the CFR was low (1%). There was a broad range of prevalent symptoms in patients, including olfactory symptoms. The majority of patients admitted had no comorbidity and the commonest comorbidity was hypertension, although it did not predict severity. Mass gathering was a risk factor for COVID-19 infection. Male gender, history of diabetes, history of lung disease, history of HIV infection, the presence of dyspnoea and fatigue were predictors of disease severity. Future studies should aim at presenting clinical and epidemiological features of COVID-19 with larger data sets.

## Data Availability

Data are available upon reasonable request to the Ministry of Public Health in Cameroon.

## Contributors

NFM and YB conceived the study. EE, HEG, and NA provided important inputs on the aims of the study. MM and JEH developed the models used in this study. PO, TP, NV, SM, FA, MO, DJ, MT and MC made important contributions regarding follow up of patients. HN, AK, GJ, EJ, ST, NSS, NNE, MNL and BL contributed to data collection. All authors contributed to writing the manuscript and approved the final version.

## Acknowledgements

We acknowledge the entire staff of the Djoungolo Hospital as well as representatives of the Ministry of Public Health.

## Patient consent for publication

Not required.

## Ethics approval

This study design and protocol were approved by the National Ethical Committee of Research for Human Health in Cameroon (Number 2020/09/1294/CE/CNERSH/SP).

## Provenance and peer review

Not commissioned; externally peer reviewed.

## Data availability statement

Data are available upon reasonable request to the Ministry of Public Health in Cameroon.

## Transparency declaration

The manuscript is an honest, accurate, and transparent account of the study being reported; no important aspects of the study have been omitted; and any discrepancies from the study as planned have been explained.

